# Achieving coordinated national immunity and cholera elimination in Haiti through vaccination

**DOI:** 10.1101/19011072

**Authors:** Elizabeth C. Lee, Dennis L. Chao, Joseph Lemaitre, Laura Matrajt, Damiano Pasetto, Javier Perez-Saez, Flavio Finger, Andrea Rinaldo, Jonathan D. Sugimoto, M. Elizabeth Halloran, Ira M. Longini, Ralph Ternier, Kenia Vissieres, Andrew S. Azman, Justin Lessler, Louise C. Ivers

**Author notes:** denotes equal contribution (Elizabeth C. Lee, Dennis L. Chao, Joseph Lemaitre, Laura Matrajt). denotes co-senior author (Andrew S. Azman, Justin Lessler, Louise C. Ivers).

## Abstract

**Background:** Cholera was introduced into Haiti in 2010. Since, there have been over 820,000 reported cases and nearly 10,000 deaths. The year 2019 has seen the lowest reported number of cases since the epidemic began. Oral cholera vaccine (OCV) is safe and effective, but has generally not been seen as a primary tool for cholera elimination due to a limited period of protection and constrained supplies. Regionally, epidemic cholera is contained to the island of Hispaniola. Hence, Haiti may represent a unique opportunity to eliminate cholera by use of OCV.

**Methods:** We assess the probability of elimination and the potential health impact of OCV use in Haiti by leveraging simulations from four independent modeling teams. For a 10-year projection period, we compared the impact of five vaccination campaign scenarios, differing in geographic scope, vaccination coverage, and rollout duration to a status quo scenario without vaccination. Teams used common calibration data and assumptions for vaccine efficacy and vaccination scenarios, but all other model features and assumptions were determined independently.

**Findings:** A two-department OCV campaign proposed in Haiti’s national plan for elimination had less than 50% probability of elimination across models, and only ambitious, nationwide campaigns had a high probability of reaching this goal. Despite their low probability of elimination, two-department campaigns averted a median of 13-58% of infections across models over the five years after the start of vaccination campaigns; a nationwide campaign implemented at the same coverage and rollout duration averted a median of 58-95% of infections across models.

**Interpretation:** Despite recent declines in cholera cases in Haiti, bold action is needed to promote elimination of cholera from the region. Large-scale cholera vaccination campaigns in Haiti offer the opportunity to synchronize nationwide immunity, providing near-term protection to the population while improvements to water and sanitation infrastructure create an environment favorable to long-term cholera elimination.

**Research in Context:** *Evidence before this study:* We searched PubMed without language or date restrictions on October 4, 2019 for all records matching (“cholera*” AND “Haiti” AND (“vaccin*” OR “elim*”)) in any field and added one known article on the probability of elimination of cholera that was not indexed by PubMed to our review. Of 94 results, four articles were not about the cholera outbreak in Haiti or the use of cholera vaccination, and 34 were not original research articles. Fourteen articles presented research on cholera biology or cholera vaccine biology, either through discussion of *Vibrio cholerae* genetics, immunogenicity of oral cholera vaccine (OCV), or prospective vaccine candidate antigens. Twenty articles assessed OCV vaccine effectiveness, evaluated OCV campaign implementation or attitudes and knowledge about cholera control, or presented lessons learned on outbreak response and policy as a result of the Haiti cholera outbreak. Seven articles were about general cholera outbreak epidemiology in Haiti, and six articles were related to cholera transmission models outside our research scope. Of the nine remaining articles, five examined the impact of potential OCV campaigns at an early time point when Haiti’s cholera outbreak still exhibited epidemic dynamics, and one other projected the impact of the OCV campaigns planned after Hurricane Matthew in 2016. Two of the articles considered prospects for cholera elimination in Haiti in 2013 and 2014 and found that further targeted interventions were needed. One final study from 2017 modeled the possibility for OCV campaigns to eliminate cholera transmission in the Ouest department within a few years.

*Added value of this study:* Previous assessments of the impact of OCV use in Haiti occurred during early points of the outbreak when OCV campaigns were unlikely to lead to cholera elimination. Our study projects cholera transmission in Haiti with multiple years of more recent data, and directly examines prospect of cholera elimination in the status quo and under various mass OCV campaign scenarios. In bringing together results from multiple modeling teams, our study provides robust evidence about the current state of cholera transmission across Haiti and the potential impact of multiple mass OCV campaign scenarios.

*Implications of all of the available evidence:* While 2019 has seen the lowest number of cholera cases in Haiti since the outbreak began, model simulations suggest that it may be possible for cholera transmission to persist without additional cholera control interventions.While a single two-department vaccination campaign may avert roughly 13-58% of infections with *V. cholerae* over a five year period, only a nationwide campaign led to a high probability of cholera elimination. Ambitious nationwide vaccination campaigns may break the cycle of endemic cholera transmission in Haiti as long-term improvements to water and sanitation infrastructure, which will limit the effects of potential re-introductions of *Vibrio cholerae*, are being made.

## Introduction

In January 2010 a massive earthquake struck Haiti, displacing over one million people and further disrupting the already inadequate water and sanitation infrastructure.^1–4^ Shortly thereafter, pandemic *Vibrio cholerae* O1 was introduced into Haiti for the first time by soldiers from the United Nations Stabilization Mission, who were themselves using deficient sanitation facilities.^5^ This confluence of events initiated one of the largest cholera outbreaks in the modern era, resulting in over 600,000 reported cases and 7,000 deaths in the first two years.^6^ Subsequent studies suggest there may have been many more deaths, especially in rural communities with limited access to health services and poor disease surveillance.^7^ Cholera has since become endemic in the country, resulting in over 820,000 reported cases and nearly 10,000 deaths as of June 2019. Although incidence rates have declined markedly in recent years, in 2018, nine of the ten departments reported a total of over 3,700 cholera cases and 41 deaths.^8^

Killed oral cholera vaccine (OCV) has become accepted as a safe and effective tool for cholera prevention and control. The standard two-dose course is 76% (95% CI 62-85%) effective against clinical disease.^9^ However, the protection from the vaccine wanes over time, and OCV is far less effective among young children than in adults (vaccine efficacy is 30% in children under five).^9^ Further, despite pre-qualification of the vaccine by the World Health Organization (WHO) in 2011, global vaccine supplies have been constrained. Approximately 23 million doses were produced (and delivered) in 2018 to serve the estimated 1.3 billion people who may be at risk worldwide.^10^ For these reasons, OCV is not generally considered a practical tool for sustained cholera elimination, as herd immunity would be difficult to achieve and maintain.

The current cholera situation in Haiti, however, may present a unique opportunity to eliminate cholera from a region through the use of vaccine as a complement to investment in long term WASH infrastructure. Outside of Haiti, the Americas are largely free from sustained cholera transmission. Only the Dominican Republic, which shares the island of Hispaniola with Haiti, has reported ongoing transmission; though at far lower rates (2,800 versus 58,700 suspected cases annually in Haiti from 2014 through 2017).^8^ It is unclear if cholera transmission could be sustained in the Dominican Republic in the absence of transmission in Haiti. Further, Haiti is a relatively small country (<13 million people) compared to other homes of endemic cholera such as the Democratic Republic of the Congo (85 million people) and Bangladesh (160 million people). If mass vaccination of the entire country could achieve coordinated cholera immunity throughout Haiti, and this immunity could be maintained for a period sufficient to flush cholera out of local water supplies, the country would become cholera-free, and would likely remain so due to the low probability of reintroduction.^11^ Elimination of cholera would also present an opportunity to focus much needed investments on transformative water and sanitation infrastructure.

Several previous studies have simulated the impact of cholera control interventions in Haiti. A few studies compared the effects of OCV campaigns and WASH improvements,^12,13^. However, these and other studies focused solely on OCV campaigns examined the impact of strategies within the first few years of the outbreak (when epidemic cholera dynamics prevailed),^12–16^ did not perform long-term projections that account for immune dynamics,^13,15,16^ or focused on a single department^17^ or historical OCV campaigns.^14,18^ Other studies considered prospects for cholera elimination in Haiti outside the context of vaccination campaigns,^19,20^ and found that additional cholera control measures were needed to achieve these goals.

In this study, we assessed the impact of five prospective oral cholera vaccination campaign scenarios as compared to a status quo scenario for a projected 10-year period in Haiti. Four independent modeling teams expanded on previously developed models of cholera transmission and vaccination interventions in Haiti^15,18,19,21–24^ to simulate the effects of mass vaccination campaigns of varying geographic scope, vaccination coverage, and rollout duration to assess the probability and time to elimination and the percentage of cases averted in each vaccination scenario. The aim of these analyses was to determine the feasibility of cholera elimination from Haiti in the status quo and through OCV use alone, and to inform ongoing policy discussions about the scope and rollout of potential OCV campaigns in Haiti in the near future.

## Methods

### Overview

We examined the health impact and feasibility of cholera elimination in Haiti with mass vaccination campaigns by establishing a consortium among four independent research teams that had previously modeled cholera transmission in Haiti. Teams fit their models to a common cholera incidence data source and generated model projections of true and reported cholera incidence 10 years past the end of the data available for model fitting. Multiple projections were produced by each model; a status quo scenario and five vaccination campaign scenarios that differed by deployment and vaccination coverage. Teams were asked to estimate the probability of cholera elimination, time to cholera elimination, and the health impact due to mass OCV campaigns for each scenario.

### Project Coordination

We discussed the goals of the project and methods with partners in the Haiti Ministry of Public Health and Population (MSPP) at the onset of this initiative for feedback on the approach and primary assumptions. Once work had started, we had multiple consultative meetings in Haiti and by teleconference with epidemiologists, researchers, and clinicians that were involved in the cholera response in Haiti in their individual capacities.

To coordinate the modeling consortium, we held bi-weekly phone calls to discuss timelines, discuss modeling progress, and troubleshoot problems. For ease of comparison, the consortium decided upon common parameters and assumptions related to vaccine protection and vaccine campaign logistics. We also shared common data sources, including weekly, department-level incidence data for model fitting,^25^ used common definitions of disease elimination, and produced comparable outputs for figures and analyses. These common assumptions are detailed below. All other modeling decisions and assumptions were left to the discretion of each team, and are described in detail in the team supplements. The final outputs for each model and model supplement underwent internal review from individuals of at least one other team.

### Data Sources

All teams calibrated their models to publicly available weekly department-level cholera reports of suspected cases from the Haiti Ministry of Public Health and Population (MSPP) website.^25^ Data were available for the epidemiological week ending on October 23, 2010 through that ending on January 12, 2019 (Figure 1). Teams also had access to optional shared data sources on diagnostic testing and prior OCV campaigns, which were used to calibrate or validate the models (Supplementary Material 1.1.1).

**Figure 1.**
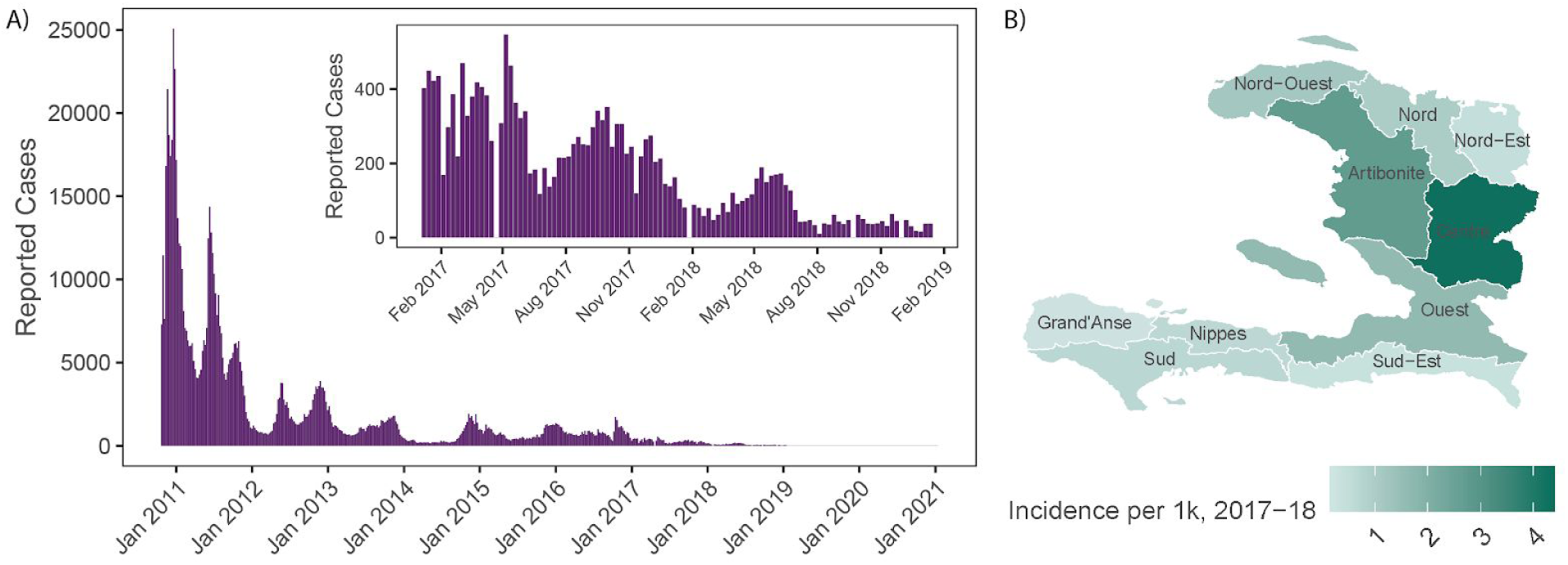
Historical cholera incidence and 10-year projections of the probability of elimination in Haiti. A) Weekly reports of cholera to the Ministère de la Santé Publique et de la Population in Haiti from October 2010 to January 2019, with an inset of the period after January 2017. The grey box represents the two-year vaccination campaign period in the modeling exercise. B) Map of reported cholera incidence per 1,000 persons from 2017-2018 across Haitian departments.

### Epidemiological Modeling

The models ranged from simple stochastic compartmental models to agent-based models of cholera dynamics in the entire country (Table 1). Model-1 (Johns Hopkins Bloomberg School of Public Health) represents all of Haiti as a single population in a stochastic compartmental model. Model-2 (Fred Hutchinson Cancer Research Center and University of Florida) and Model-3 (École Polytechnique Fédérale de Lausanne) are, respectively, deterministic and stochastic metapopulation models, each with an independent approach to modeling inter-departmental connectivity and the dynamics of cholera reservoirs. Model-4 (Institute for Disease Modeling) is an agent based model that uses a synthetic representation of the Haitian population, its household structure, connectivity, and interaction with aquatic reservoirs. All modeling teams have comprehensive supplementary method, results, and model code (See Supplementary Material Sections 2.1, 3.1, 4.1, and 5.1), which are assembled collectively at the summary DOI: 10.5281/zenodo.3361800.

**Table 1.** Summary of key model features across teams.

| **Model (refs)** | **Spatial Scale** | **Seasonality Function** | **Environmental Transmission** | **Age Structure** | **Spatial Transmission Dynamics** |
| --- | --- | --- | --- | --- | --- |
| 1<sup>15</sup> | National | Basis splines | None | None | None |
| 2 | Department | Sinusoidal | Yes | None | Road and river networks |
| 3<sup>18,19,22-24</sup> | Department | Rainfall-driven | Yes | None | Calibrated human mobility |
| 4<sup>16</sup> | 1 km x 1 km grid | Rainfall-driven | Yes | Yes | Road and river networks, commuting |

Teams simulated six scenarios that employed different combinations of parameters for vaccine campaign logistics and vaccination coverage. The parameters are described in detail below sections and the six scenarios are summarized as follows (Figure 2):

1. Status quo (no vaccination)
2. Two-department OCV campaign over 2 years with baseline vaccination coverage (2-department)
3. Three-department OCV campaign over 2 years with baseline vaccination coverage (3-department)
4. National OCV campaign over 2 years with baseline vaccination coverage (national)
5. National OCV campaign over 5 years with baseline vaccination coverage (slow national)
6. National OCV campaign over 2 years with high vaccination coverage (high coverage national)

**Figure 2.**
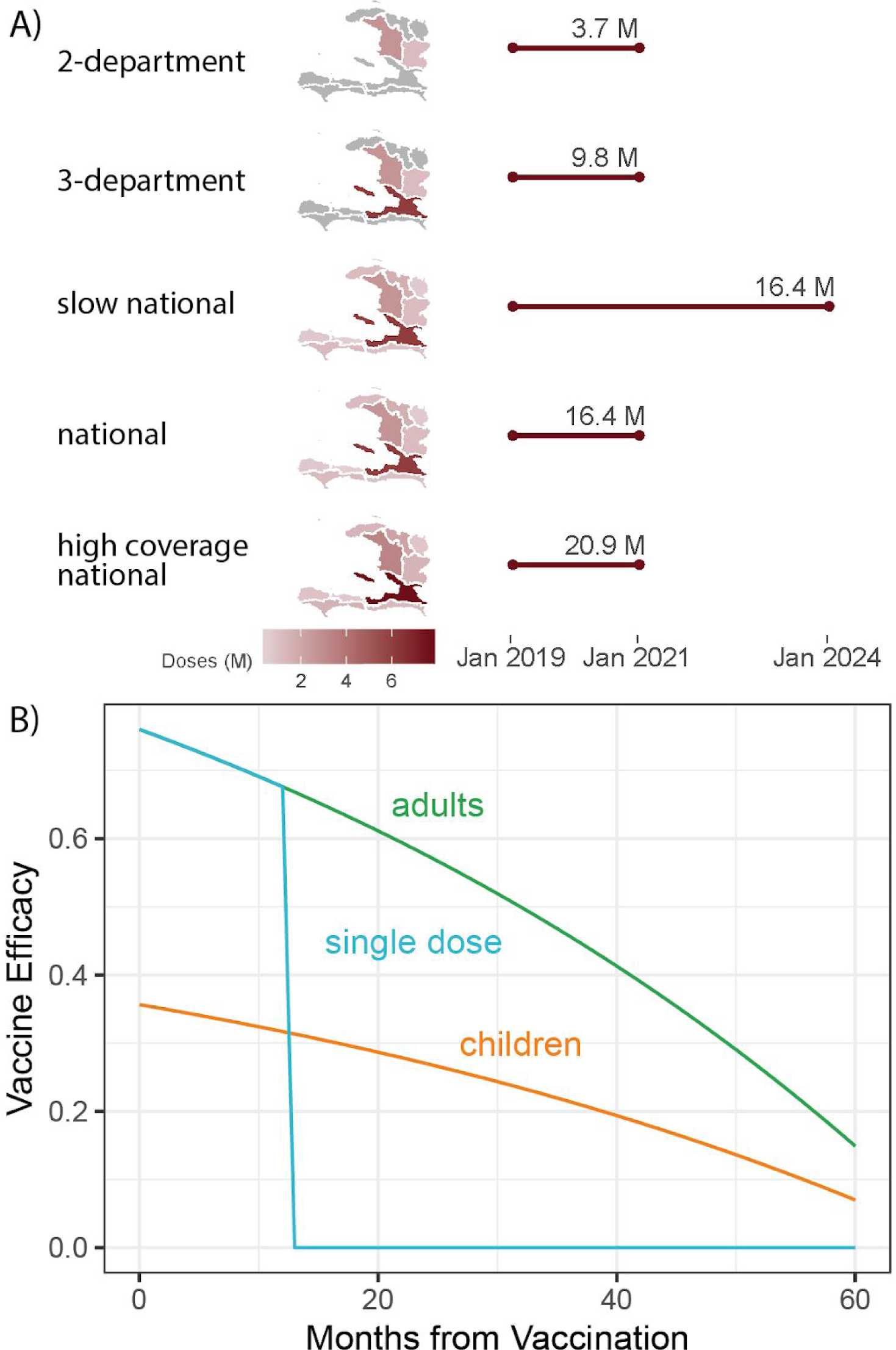
Vaccination campaign scenarios and vaccine efficacy assumptions. A) Summary of the geographic scope, vaccination campaign deployment, and total number of OCV doses needed for each of the five modeled vaccination campaign scenarios. B) Common vaccine efficacy assumptions for 2-doses administered to adults, children, and 1-dose.

### Vaccine Campaign Logistics

We implemented four vaccine campaign deployment strategies, each starting the day or week (depending on model implementation) after the last data point used for model calibration (week ending January 12, 2019). Campaigns targeted departments in order of 2017-2018 cumulative incidence from highest to lowest (Figure 1, Table S1).

### The campaign deployment strategies are as follows

- Two-department: Vaccination limited to the departments of Centre and Artibonite over a 2-year period, similar to plans outlined in the national cholera elimination plan for Haiti^26^
- Three-department: Vaccination limited to the departments of Centre, Artibonite, and Ouest (which includes the populous capital Port-au-Prince) over a 2-year period.
- National: Countrywide vaccination implemented over a 2-year period.
- Slow national: Countrywide vaccination implemented over a 5-year period.

### Vaccination Coverage

Killed OCVs are licensed as a two-dose regimen, with doses taken at least two weeks apart.^9^ All simulated campaigns aimed to vaccinate everyone with two doses, however, following data from previous vaccination campaigns, a fraction of individuals only received a single dose and some remained unvaccinated. Our baseline scenario assumes that vaccine coverage is the same in all departments with 70% two-dose coverage, 10% one-dose coverage and 20% receiving no vaccine. We also simulated one ‘high coverage’ campaign, where departments achieved 95% two-dose coverage, 1.67% one-dose coverage and 3.33% unvaccinated at the end of the campaign.

### Vaccine Efficacy Among Adults

All teams assumed that initial vaccine efficacy was 76%, as estimated by a recent case-control study in Haiti.^27^ We estimated waning vaccine efficacy for 60 months after vaccination by fitting a log-linear weighted regression model to the raw data from a published meta-analysis on killed OCV efficacy (excluding a single outlier estimate of vaccine efficacy in India after 5 years to be conservative) (Figure 2, Table S3).^9^ We assumed that the vaccine afforded no protection after the end of five years.

### Vaccine Efficacy Among Children

Following estimates from a recent meta-analysis, the average efficacy among children under five years old was 46.9% (0.30/0.64) as effective as that in adults.^9^ As there are limited data on vaccine efficacy among children, we used this ratio as a conservative multiplier to adjust the adult vaccine efficacy for vaccine efficacy among children under five (Figure 2, Table S3).

### Vaccine Efficacy from a Single Dose

In the first year after vaccination, individuals who received a single vaccine dose were assumed to have the same protection as those with two doses, after which the single dose efficacy dropped to zero (Figure 2, Table S3).^9,27–29^

### Model Outcomes

Each team estimated the “probability of elimination” within ten years after the start of vaccination campaigns, which was defined as the proportion of simulations that achieved less than one infection with *Vibrio cholerae* (includes reported and unreported infections) over at least 52 consecutive weeks. The ten-year period without resurgence was deemed adequate to limit the possibility of cholera reseeding from human or environmental reservoirs. As defined in these experiments, elimination represents a state of *“no underlying transmission,”* and not a state of *“no reported cases*.*”* We also recorded the “elimination date” for each simulation, which was defined as the first date where a simulation achieved less than one cholera infection for at least 52 consecutive weeks.

In addition to elimination metrics, we calculated the number and percentage of infections averted in vaccination campaign scenarios after the start of vaccination campaigns (as compared to the status quo scenario).

## Results

Teams performed model selection, model calibration, and assessment of model fit independently, as described in the Supplementary Material of each individual team supplement. Model projections for true infections with *Vibrio cholerae* and reported cholera cases are available for each scenario in Figures S1-S12.

The consensus across the four models is that a two-year nationwide campaign with coverage similar to that achieved by previous, smaller scale, OCV campaigns in Haiti (70% two-dose coverage)^30,31^ has a moderate chance of achieving true cholera elimination at five years post-campaign (34-100% of simulations, summarized across teams) (Figure 3, Figure S13, Table S4). If high coverage is achieved (95% two-dose coverage), the models agree that cholera elimination is almost guaranteed after a nationwide campaign (88-100% of simulations). In contrast, simulations with vaccine deployments based on Haiti’s current strategy to eliminate cholera, which aims to target 1.8 million people primarily in the two most cholera-affected departments (Centre and Artibonite),^32^ suggest there is a very low probability for this strategy to achieve elimination (0-33% of simulations) through the impact of OCV alone. There was a substantial difference between the outcomes of the 2-department and 3-department campaigns because it administered over 2.5 times more doses; the third highest incidence department (Ouest) is the most populous department in Haiti. While models were designed primarily to examine the impact of vaccination campaigns, we also projected cholera incidence in the status quo, without added vaccination campaigns, and found that there was a 0-18% probability of elimination by January 2024.

**Figure 3.**
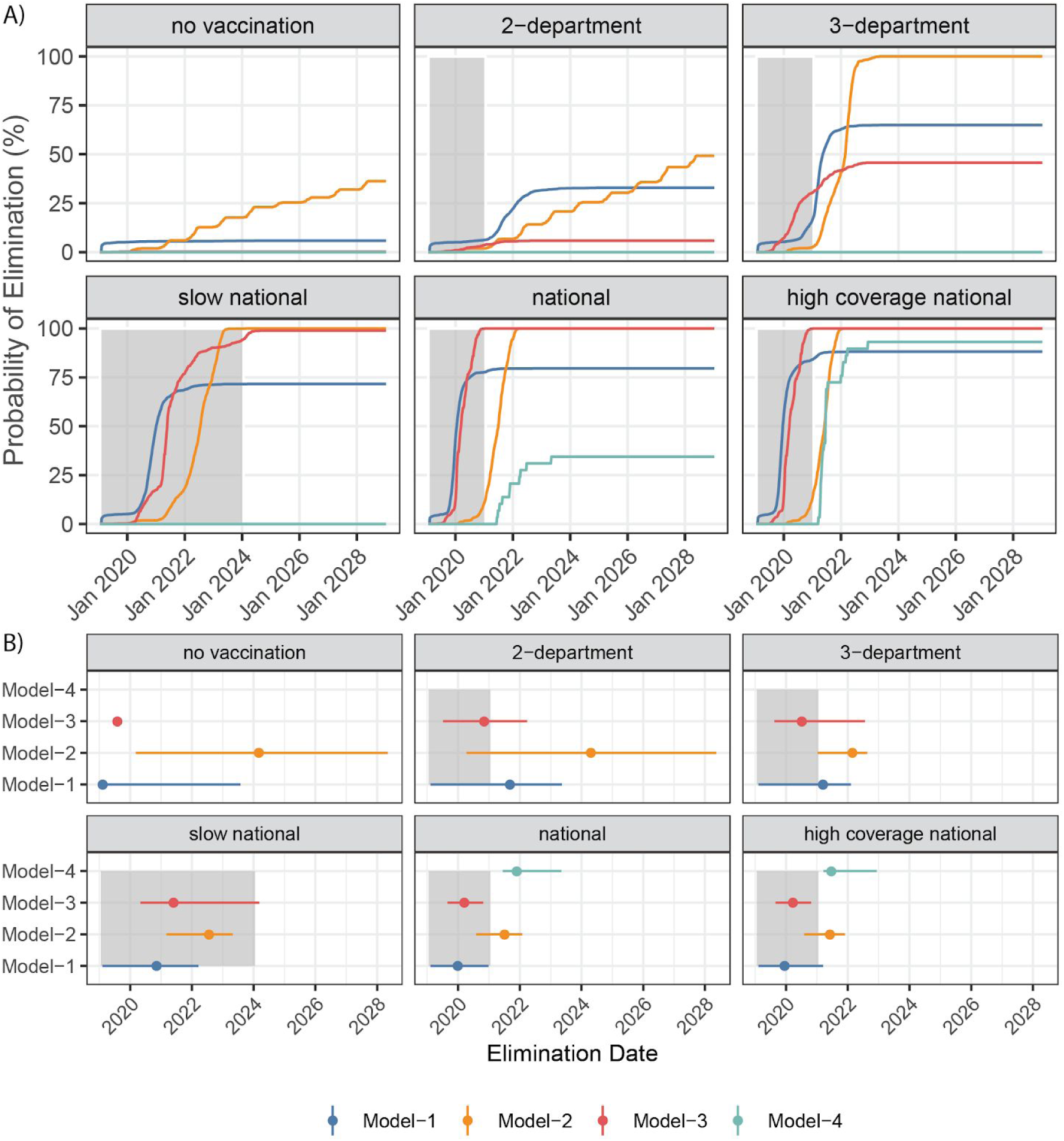
Model outcomes for probability of elimination and elimination date. A) Probability of elimination across simulations during the 10-year projection period across four models (colors) in six primary scenarios (sub-panels). B) Median elimination date (and 95% CI) for each model (colors) and scenario (sub-panels) across simulations that achieved elimination. Model scenarios with no depicted information had zero probability of elimination. For each sub-panel, the grey shaded area depicts the duration of the vaccination campaign.

We examined the date of elimination (first date among at least 52 consecutive weeks where there was less than one infection) among model and scenario simulations that achieved elimination (Figure 3). There was substantial heterogeneity in when simulations achieved elimination; for most vaccination scenarios, median elimination dates across models fell roughly within a 2-year window. The median elimination date for Model-1 was within one year of the end of vaccination rollout for the 2- and 3-department campaigns (September and March 2021, respectively) and in the middle of the vaccination rollout for all three national campaigns (ranging from December 2019 to November 2020). The median elimination date for Model-2 was mostly outside of the vaccination rollout period of campaigns (ranging from June 2021 to April 2024), while the median elimination date for Model-3 was always achieved before the end of the vaccination campaigns (ranging from March 2020 to May 2021). Model-4 only achieved elimination in the national and high coverage national scenarios with median elimination dates in November 2021 and June 2021, respectively.

Beyond assessing the feasibility of cholera elimination through vaccination, we compared the percentage of averted infections within five years of the start of vaccination campaigns for various scenarios (Figure 4, Figure S14). The 2-department campaign averted a median of 13-58% of infections, while the national campaign averted a median of 58-94% of infections across models.

**Figure 4.**
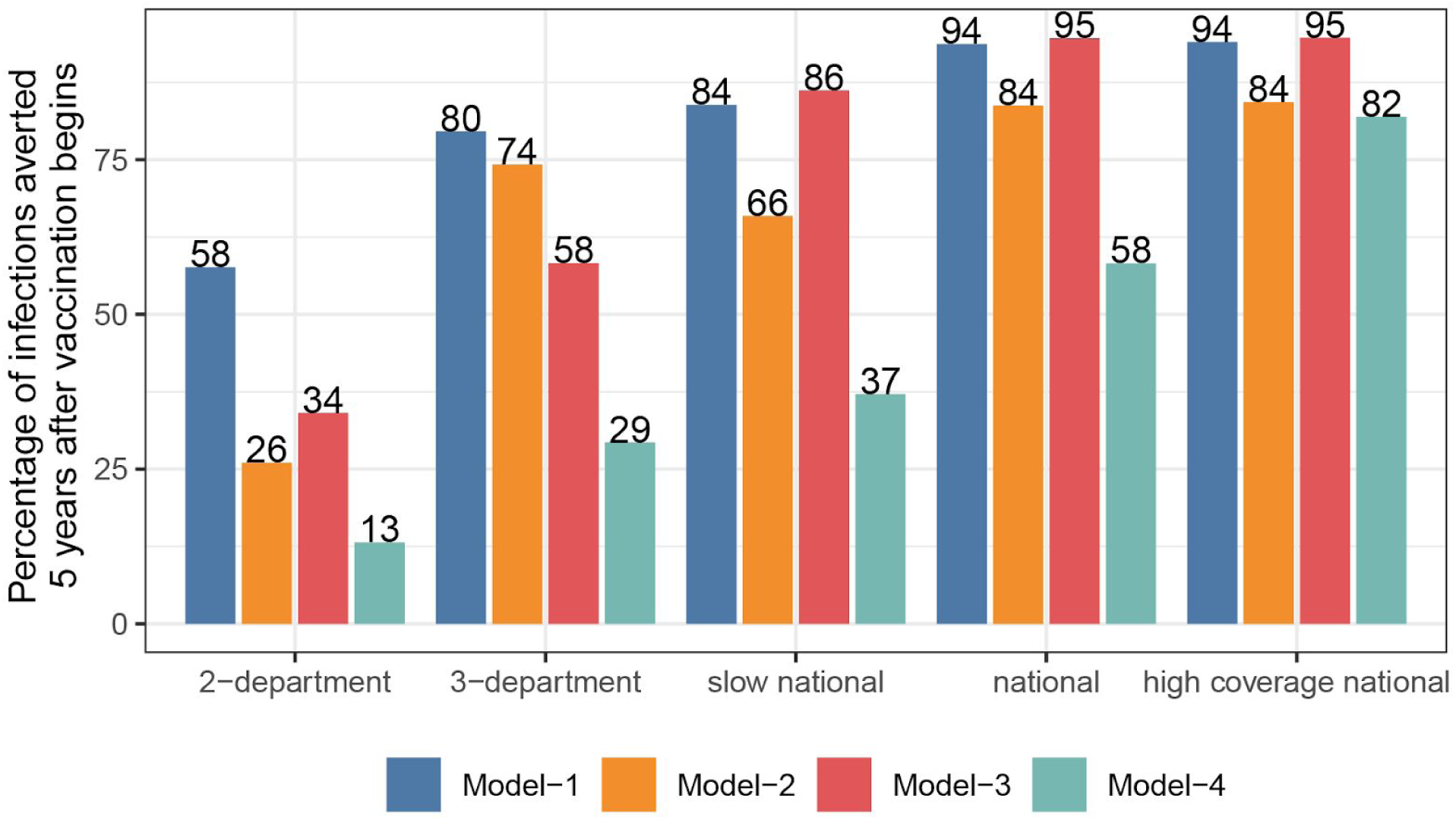
Model outcomes for percentage of infections averted. Median percentage of infections averted within five years after the start of vaccination campaigns, by model and vaccination campaign scenario.

## Discussion

While the cholera epidemic has declined substantially in Haiti over the past year, our study suggests that cholera transmission may persist in the face of low incidence at historically-observed rates of underreporting and asymptomatic infection. The most recent national plan for the elimination of cholera in Haiti proposes to target the two departments with the highest cholera incidence (Centre and Artibonite),^32^ and our multimodeling effort suggests that this strategy may avert 13-58% of infections within a five year period and yield a low probability (0-33%) of achieving elimination. Only when the models simulated a nationwide vaccination campaign with relatively short rollout duration (two years) did all models project at least some probability of elimination with 58-95% of infections averted within a five year period.

Our study lends credence to the idea that cholera elimination in Haiti through mass vaccination is an achievable goal, albeit one that may require a larger and more ambitious campaign than those previously achieved in the country. As the United Nations acknowledges responsibility for the introduction of cholera into Haiti, support for such an ambitious effort would be a direct way to start making amends for this tragedy. Even if unsuccessful in eliminating cholera, such a campaign would result in reductions of cholera incidence in Haiti, providing substantial, if not sustained, benefits to its inhabitants.

The call for mass OCV use in Haiti should in no way be construed as a call to decrease efforts to improve access to safely managed and sustainable water and sanitation facilities in the country, and instead should be used to leverage larger funding for this critical infrastructure.

Water and sanitation improvements protect individuals from a suite of diseases beyond cholera, and universal access to clean water and sanitation is a critical component of the United Nations Sustainable Development Goals. Further, until there is widespread access to clean water and adequate sanitation, Haiti remains vulnerable to reintroduction of cholera. However, halting endemic transmission through vaccination could remove cholera from the Americas for decades, protecting vulnerable populations from one of the deadliest water-borne pathogens.

This is perhaps all the more relevant in the current sociopolitical context of Haiti in which reports of water insecurity in urban areas abound as fuel crises and political protests hamper access to clean water for some of the population.^33^

Cholera incidence in Haiti has been steadily declining since 2012, and there is some thought that the country may be on track to elimination with current control activities. Despite this decline in reported cases, model simulations suggest the probability of elimination (in contrast to the absence of clinical cases) is low (0-18% of simulations) without changes to current conditions.

Recent resurgences of measles in Madagascar^34^ and elsewhere show that we should be cautious about the sustainability of apparent reductions in disease incidence. As immunity from infection and previous vaccine campaigns wanes in Haiti, it is not unreasonable to believe the country may soon be at risk for a cholera resurgence. Our modeling shows that it is possible to have periods of low to no *detected* cholera cases followed by disease resurgence (Figures S7-S12).

Individual models each had their own set of limitations with regard to its mechanisms for characterizing cholera transmission and vaccine dynamics; one or multiple models in our exercise did not include spatial heterogeneity in cholera transmission, population movement data, population dynamics, differences in vaccine efficacy by age, and environmental reservoir or transmission components (See Supplementary Material). To combat these individual model deficiencies, we interpreted model results collectively and focused on the convergent findings of model outcomes. The collective exercise was limited more generally by our lack of information related to community-wide rates of the loss of acquired immunity, the importance of environmental reservoirs in cholera transmission, and reporting and asymptomatic rates of cholera in Haiti.

If vaccine supplies and other resources were unconstrained, there would be little downside to a mass vaccination campaign (the vaccine has few if any side effects). In reality, cholera vaccine supplies are severely limited and resources for public health are insufficient. Haiti’s unique situation provides a rare opportunity to use OCV to eliminate cholera from an entire region of the world, rather than as a bandaid to respond to continuing flare ups and disasters. Our modeling studies suggest that this goal is achievable with a high quality, large-scale campaign with high population coverage. Such an effort would represent an innovative, and perhaps radical, use of public health resources, but might offer substantial long-term benefits and a reprieve to the already-stressed public health facilities and emergency response sector in Haiti. If a national OCV campaign prevented future outbreaks for decades or contributed to the elimination of cholera from the entire region, it would ultimately consume fewer resources than the status quo. These resources could then be devoted to transformative systems change in the water and sanitation sector, accelerating progress towards achieving Sustainable Development Goal 6.

To take this bold approach, the global health community, from donors to local actors, needs to step up. The United Nations Haiti Cholera Response Multi-Partner Trust Fund, established in response to the United Nations’ role in cholera’s introduction into Haiti, is woefully short of its 400-million-dollar fundraising goal. Haiti’s cholera control plan is likewise underfunded. Now that cases of cholera are at an all-time low in Haiti, a national OCV campaign could serve as a rallying point for local and international stakeholders; potentially eliminating an illness that has brought almost a decade of suffering to the poorest and making a first step in protecting the country from water-borne diseases over the long haul.

## Data Availability

All teams calibrated their models to publicly available weekly department-level cholera reports of suspected cases from the Haiti Ministry of Public Health and Population (MSPP) website. All modeling teams have provided a detailed supplementary methods and results section, which are assembled collectively
at the summary DOI: 10.5281/zenodo.3361800.

https://doi.org/10.5281/zenodo.3362554

## Acknowledgments

The authors are grateful for the input and critical discussions with various public health officials acting in their individual capacities.

## References

1 Lantagne D, Balakrish Nair G, Lanata CF, Cravioto A. The Cholera Outbreak in Haiti: Where and How did it begin? Berlin: Springer, 2013.

2 Walton DA, Ivers LC. Responding to cholera in post-earthquake Haiti. N Engl J Med 2011; 364 : 3–5.

3 Jenson D, Szabo V, Duke FHI Haiti Humanities Laboratory Student Research Team. Cholera in Haiti and other Caribbean regions, 19th century. Emerg Infect Dis 2011; 17 : 2130–5.

4 Gelting R, Bliss K, Patrick M, Lockhart G, Handzel T. Water, sanitation and hygiene in Haiti: past, present, and future. Am J Trop Med Hyg 2013; 89 : 665–70.

5 Frerichs RR, Keim PS, Barrais R, Piarroux R. Nepalese origin of cholera epidemic in Haiti. Clin Microbiol Infect 2012; 18 : E158–63.

6 Barzilay EJ, Schaad N, Magloire R, et al. Cholera surveillance during the Haiti epidemic--the first 2 years. N Engl J Med 2013; 368 : 599–609.

7 Luquero FJ, Rondy M, Boncy J, et al. Mortality Rates during Cholera Epidemic, Haiti, 2010-2011. Emerg Infect Dis 2016; 22 : 410–6.

8 World Health Organization. Weekly Epidemiological Record. https://www.who.int/cholera/statistics/en/.

9 Bi Q, Ferreras E, Pezzoli L, et al. Protection against cholera from killed whole-cell oral cholera vaccines: a systematic review and meta-analysis. Lancet Infect Dis 2017; 17 : 1080–8.

10 Ali M, Nelson AR, Lopez AL, Sack DA. Updated global burden of cholera in endemic countries. PLoS Negl Trop Dis 2015; 9 : e0003832.

11 Lewnard JA, Antillón M, Gonsalves G, Miller AM, Ko AI, Pitzer VE. Strategies to Prevent Cholera Introduction during International Personnel Deployments: A Computational Modeling Analysis Based on the 2010 Haiti Outbreak. PLoS Med 2016; 13 : e1001947.

12 Fung IC-H, Fitter DL, Borse RH, Meltzer MI, Tappero JW. Modeling the effect of water, sanitation, and hygiene and oral cholera vaccine implementation in Haiti. Am J Trop Med Hyg 2013; 89 : 633–40.

13 Andrews JR, Basu S. Transmission dynamics and control of cholera in Haiti: an epidemic model. Lancet 2011; 377 : 1248–55.

14 Tuite AR, Tien J, Eisenberg M, Earn DJD, Ma J, Fisman DN. Cholera epidemic in Haiti, 2010: using a transmission model to explain spatial spread of disease and identify optimal control interventions. Ann Intern Med 2011; 154 : 593–601.

15 Azman AS, Luquero FJ, Ciglenecki I, Grais RF, Sack DA, Lessler J. The Impact of a One-Dose versus Two-Dose Oral Cholera Vaccine Regimen in Outbreak Settings: A Modeling Study. PLoS Med 2015; 12 : e1001867.

16 Chao DL, Halloran ME, Longini IM Jr. Vaccination strategies for epidemic cholera in Haiti with implications for the developing world. Proc Natl Acad Sci U S A 2011; 108 : 7081–5.

17 Kirpich A, Weppelmann TA, Yang Y, Morris JG Jr, Longini IM Jr. Controlling cholera in the Ouest Department of Haiti using oral vaccines. PLoS Negl Trop Dis 2017; 11 : e0005482.

18 Pasetto D, Finger F, Camacho A, et al. Near real-time forecasting for cholera decision making in Haiti after Hurricane Matthew. PLoS Comput Biol 2018; 14 : e1006127.

19 Bertuzzo E, Finger F, Mari L, Gatto M, Rinaldo A. On the probability of extinction of the Haiti cholera epidemic. Stochastic Environmental Research and Risk Assessment. 2016; 30 : 2043–55.

20 Rebaudet S, Gazin P, Barrais R, et al. The dry season in haiti: a window of opportunity to eliminate cholera. PLoS Curr 2013; 5. DOI: 10.1371/currents.outbreaks.2193a0ec4401d9526203af12e5024ddc.

21 Chao DL, Longini IM, Glenn Morris J. Modeling Cholera Outbreaks. Cholera Outbreaks. 2013; : 195–209.

22 Lemaitre J, Pasetto D, Perez-Saez J, Sciarra C, Wamala JF, Rinaldo A. Rainfall as a driver of epidemic cholera: Comparative model assessments of the effect of intra-seasonal precipitation events. Acta Trop 2019; 190 : 235–43.

23 Rinaldo A, Bertuzzo E, Mari L, et al. Reassessment of the 2010-2011 Haiti cholera outbreak and rainfall-driven multiseason projections. Proc Natl Acad Sci U S A 2012; 109 : 6602–7.

24 Bertuzzo E, Mari L, Righetto L, et al. Prediction of the spatial evolution and effects of control measures for the unfolding Haiti cholera outbreak. Geophysical Research Letters. 2011; 38. DOI: 10.1029/2011gl046823.

25 Profil statistique du Cholera. Republique d’Haiti, Ministère de la Santé Publique et de la Population. http://mspp.gouv.ht/newsite/.

26 Ministere de la Sante Publique et de la Population et Direction Nationale de L’Eau Potable et de L’Assainissement RD. Plan National D’Elimination Du Cholera, Developpement Du Moyen Terme, Juillet 2016 - Decembre 2018. 2016.

27 Franke MF, Ternier R, Jerome JG, Matias WR, Harris JB, Ivers LC. Long-term effectiveness of one and two doses of a killed, bivalent, whole-cell oral cholera vaccine in Haiti: an extended case-control study. Lancet Glob Health 2018; 6 : e1028–35.

28 Ferreras E, Chizema-Kawesha E, Blake A, et al. Single-Dose Cholera Vaccine in Response to an Outbreak in Zambia. N Engl J Med 2018; 378 : 577–9.

29 Qadri F, Ali M, Lynch J, et al. Efficacy of a single-dose regimen of inactivated whole-cell oral cholera vaccine: results from 2 years of follow-up of a randomised trial. Lancet Infect Dis 2018; 18 : 666–74.

30 Tohme RA, François J, Wannemuehler K, et al. Oral Cholera Vaccine Coverage, Barriers to Vaccination, and Adverse Events following Vaccination, Haiti, 2013. Emerg Infect Dis 2015; 21 : 984–91.

31 Pierre K. Use and perspectives of the oral cholera vaccine (OCV) as a component of the cholera response. December 5–6, 2018. https://www.fondation-merieux.org/en/events/5th-gtfcc-working-group-on-oral-cholera-vaccine/.

32 Plan national d’elimination du cholera, Developpement du plan long terme 2018-2022. Ministère de la Santé Publique et de la Population, Direction Nationale de L’eau Potable et de L’assainissement, Republic of Haiti, 2019. http://mspp.gouv.ht/newsite/.

33 Thornton C, Dupain E, Barnes T, Castillo J. Humanitarian crisis increases in Haiti as anti-government protests grip the nation. CNN. 2019; published online Oct 3. https://www.cnn.com/2019/10/03/americas/haiti-anti-government-protests/index.html.

34 WHO | Measles – Madagascar. 2019; published online Jan 25. http://www.who.int/csr/don/17-january-2019-measles-madagascar/en/ (accessed May 14, 2019).

